# Deep learning approach for automatic assessment of schizophrenia and bipolar disorder in patients using R-R intervals

**DOI:** 10.1101/2025.03.25.25324600

**Authors:** Kamil Książek, Wilhelm Masarczyk, Przemysław Głomb, Michał Romaszewski, Krisztián Buza, Przemysław Sekuła, Michał Cholewa, Katarzyna Kołodziej, Piotr Gorczyca, Magdalena Piegza

**Affiliations:** Faculty of Mathematics and Computer Science, Jagiellonian University Łojasiewicza 6, 30-348 Kraków, Poland; Institute of Theoretical and Applied Informatics, Polish Academy of Sciences Bałtycka 5, 44-100 Gliwice, Poland; Department of Psychiatry, Faculty of Medical Sciences in Zabrze, Medical University of Silesia, Pyskowicka 49, 42-612 Tarnowskie Góry, Poland; Faculty of Finance and Accountancy, Budapest Business University, Buzogány utca 10-12, 1149 Budapest, Hungary; BioIntelligence Group, Department of Mathematics-Informatics, Sapientia Hungarian University of Transylvania, Targu Mures, Romania

## Abstract

Schizophrenia and bipolar disorder are severe mental illnesses that significantly impact quality of life. These disorders are associated with autonomic nervous system dysfunction, which can be assessed through heart activity analysis. Heart rate variability (HRV) has shown promise as a potential biomarker for diagnostic support and early screening of those conditions. This study aims to develop and evaluate an automated classification method for schizophrenia and bipolar disorder using short-duration electrocardiogram (ECG) signals recorded with a low-cost wearable device.

We conducted classification experiments using machine learning techniques to analyze R-R interval windows extracted from short ECG recordings. The study included 60 participants – 30 individuals diagnosed with schizophrenia or bipolar disorder and 30 control subjects. We evaluated multiple machine learning models, including Support Vector Machines, XGBoost, multilayer perceptrons, Gated Recurrent Units, and ensemble methods. Two time window lengths (about 1 and 5 minutes) were evaluated. Performance was assessed using 5-fold cross-validation and leave-one-out cross-validation, with hyperparameter optimization and patient-level classification based on individual window decisions. Our method achieved classification accuracy of 83% for the 5-fold cross-validation and 80% for the leave-one-out scenario. Despite the complexity of our scenario, which mirrors real-world clinical settings, the proposed approach yielded performance comparable to advanced diagnostic methods reported in the literature.

The results highlight the potential of short-duration HRV analysis as a cost-effective and accessible tool for aiding in the diagnosis of schizophrenia and bipolar disorder. Our findings support the feasibility of using wearable ECG devices and machine learning-based classification for psychiatric screening, paving the way for further research and clinical applications.

**Author summary:** Mental health conditions such as schizophrenia and bipolar disorder can be challenging to diagnose, but early detection is crucial to better outcomes. In our study, we explored a simple but powerful idea: using heart rate variability (HRV), the natural fluctuations in time between heartbeats, as a potential indicator of these disorders. As HRV reflects the balance of the autonomic nervous system, research has shown that people with schizophrenia and bipolar disorder often have distinct HRV patterns, suggesting that heart activity analysis could support mental health screening. We tested whether a wearable heart monitor, similar to a fitness tracker, could help recognize these patterns using artificial intelligence. By analyzing heart activity records from 60 individuals, some with these conditions and others without, we trained several machine learning models to differentiate between them. Our approach successfully classified individuals with about 80% accuracy. Although not a replacement for clinical diagnosis, heart monitoring combined with artificial intelligence could help detect signs of mental illness, leading to faster interventions. Our work opens the door for further studies and, ultimately, the development of easy-to-use tools that could support doctors and improve patient care.

## 1 Introduction

Schizophrenia and bipolar disorder (BD) are both severe, chronic mental disorders with a high population prevalence rate of more than 1% [1] and a well-documented increasing tendency in the case of schizophrenia [**?**, 2]. Both diseases have a significant negative impact on patients’ quality of life [2], including social interactions, employment levels and income [3]. Both diseases overlap in symptoms and pathophysiology and can be discussed in terms of spectrum and continuity [4]. Diagnosis of the disease in its early, prodromal stage, when treatment is most effective, is challenging [5]. Diagnosis usually occurs only when symptoms become more pronounced and aggravation of symptoms in these two diseases is one of the main reasons for psychiatric ward admission [6]. This increases treatment costs and the burden placed on the mental healthcare system [7]. Thus, early diagnosis and intervention are crucial for the improvement of treatment costs and outcomes [8]. Current diagnostic methods are based purely on clinical interviews by trained physicians [9, 10]. It is time- and resource-consuming and open to debate whether this approach is truly objective [11, 12].

The clinical diagnosis may change over time, with a retrospective consistency (a concept similar to sensitivity) of 91.5% and 89.3% in a clinical setting for schizophrenia and BD, respectively. In addition, the consistency of the diagnosis is significantly different when the patients are assessed in psychiatric emergency and out-patient settings, with a retrospective consistency lower than 70% [13]. Although the results in a clinical setting could be considered satisfactory and provide a workable baseline, there is a desire to improve accuracy and achieve even better outcomes. More importantly, the findings outside the clinical setting highlight the importance of improving the overall quality of diagnostics, regardless of location, using accessible and user-friendly on-site diagnostic tools.

Various sources of biological data are taken into consideration to enhance and objectify the psychiatric diagnostic process. Among them, broad-ranging approaches, including MRI [14, 15], EEG [16, 17], fMRI [18, 19], genetic analysis [20], and autonomic nervous system (ANS)-mediated physiological patterns such as R-R interval derived heart rate variability (HRV) [21] offer a rich source of data on the functioning and structure of the brain. This complexity and richness of data pose a challenge in itself, e.g. in cases such as MRI and genetic profiling, as these types of data have a multidimensional character [22, 23]. This can lead to a significantly larger number of false associations due to a higher degree of confounding effects [24] and makes it challenging and resource-intensive to identify the desired features [25, 26]. Most importantly, global economic reality limits the access of most people to clinical settings with expensive equipment and trained personnel and consequently, to effective psychiatric care [27, 28].

A possible solution to this problem might be the use of a diagnostic approach based on physiological biomarkers such as physical activity [29], galvanic skin response [30], pupillary response [31] or HRV [32]. These biomarkers can be easily obtained through currently available wearable devices [33–35], fairly inexpensive technology that can be used even outside the clinical setting [36]. Among these biomarkers, HRV stands out as particularly promising. Both schizophrenia and BD share numerous common genetic, endophenotypic, and phenomenological traits [37, 38]. One such particular trait is an ANS dysregulation that can be observed through changes in HRV [21, 32]. The classification of HRV patterns in a given individual could provide an objective source of symptom information within the diagnostic process [32, 39, 40]. A robust wearable device-based classification could significantly support the low-cost, widespread early diagnosis of schizophrenia and BD.

HRV-based wearable diagnostic systems have received significant attention in recent years. The authors of [33] propose a Mobile Health (mHealth) wearable device, which collects HRV, electrodermal activity (EDA) and movement data. After six days of wearing, the recorded data showed a statistically significant difference in HRV values between participants with schizophrenia and healthy controls, confirming previous results. However, this analysis was limited to long recording windows (*>* 8h) and did not consider the classification accuracy in a diagnostic setting. A similar approach is used by [41], where a disposable adhesive patch HR and activity sensor worn for several days are used to classify whether a patient is diagnosed with schizophrenia or belongs to a healthy control. This approach has achieved a promising AUC (area under a receiver operating characteristic curve) score of 0.96 for eight-day windows and 0.91 for two-day windows. However, this method is possibly limited by the manual choice of features, the lack of hyperparameter optimization for the classifier, and is restricted to a minimum of two days of patient observation.

A different approach is used by [42], which uses short time frame ECG measurements (25-minute windows), together with structured yoga exercises. The classification, based on the thresholding Z-score computed from Linear Discriminant Analysis (LDA), achieves a promising score of sensitivity of 91% and specificity of 81%. However, the sample size is limited (only 12 participants with schizophrenia) and, possibly because of that, no cross-validation protocol is mentioned, which severely limits the generalization of the reported scores. A wearable HRV-based setup was also successfully tested for hallucination spectrum experiences (HSE) and paranoia assessment [43], for the detection of stress in working people [44], as well as for the monitoring of symptoms in early psychosis [45] and anxiety disorders [46].

In this article, we study a classification approach for short-duration R-R interval windows to differentiate the data as belonging to a patient with schizophrenia / BD or a control group. R-R intervals are directly extracted from the ECG signal and various HRV metrics are based on their values [39, 47]. Therefore, R-R intervals are a rich source of knowledge about ANS functioning and their consecutive values can be used as input features for classifiers.

Our main contribution is the assessment of the accuracy of a wearable-recorded classification-based diagnosis protocol. In reference to previous works, our specific contributions are as follows:

1. Larger data sample analyzed (60 participants vs 24/28 in previous works reporting classification results), distributed in a publicly available dataset;
2. State-of-the-art classification test scheme, including two levels of cross-validation, allowing for unbiased patient evaluation with automatic methods’ hyperparameter optimization;
3. Comparison of several state-of-the-art machine learning classification approaches, including deep neural networks, allowing for their comprehensive evaluation, without hand-picked features.

Our approach can therefore be viewed as method-independent, providing a reference assessment that directly corresponds to a real-world clinical setting. This is particularly important as recent studies emphasize the need for advanced, automated ECG analysis techniques in various clinical applications [48]. The accuracy obtained (83% in the 5-fold cross-validation scenario and 80% in the leave-one-out cross-validation scenario) provides a solid, promising result for further applications. We view the application of our work as a prospect for the development of diagnostic and disease monitoring tools that combine traditional disease criteria based on observable symptoms [9, 10] with physiological parameters such as the ECG signal for more accurate diagnosis [49]. Furthermore, we hope that in the future, with the implementation of biological marker-based machine learning-supported diagnostic methods, it will be possible to enhance predictive validity to assess the possibility of schizophrenia or BD in their prodromal state [50].

The outline of our method is presented in Fig. 1 and will be discussed in detail in the next section.

**Fig 1.**
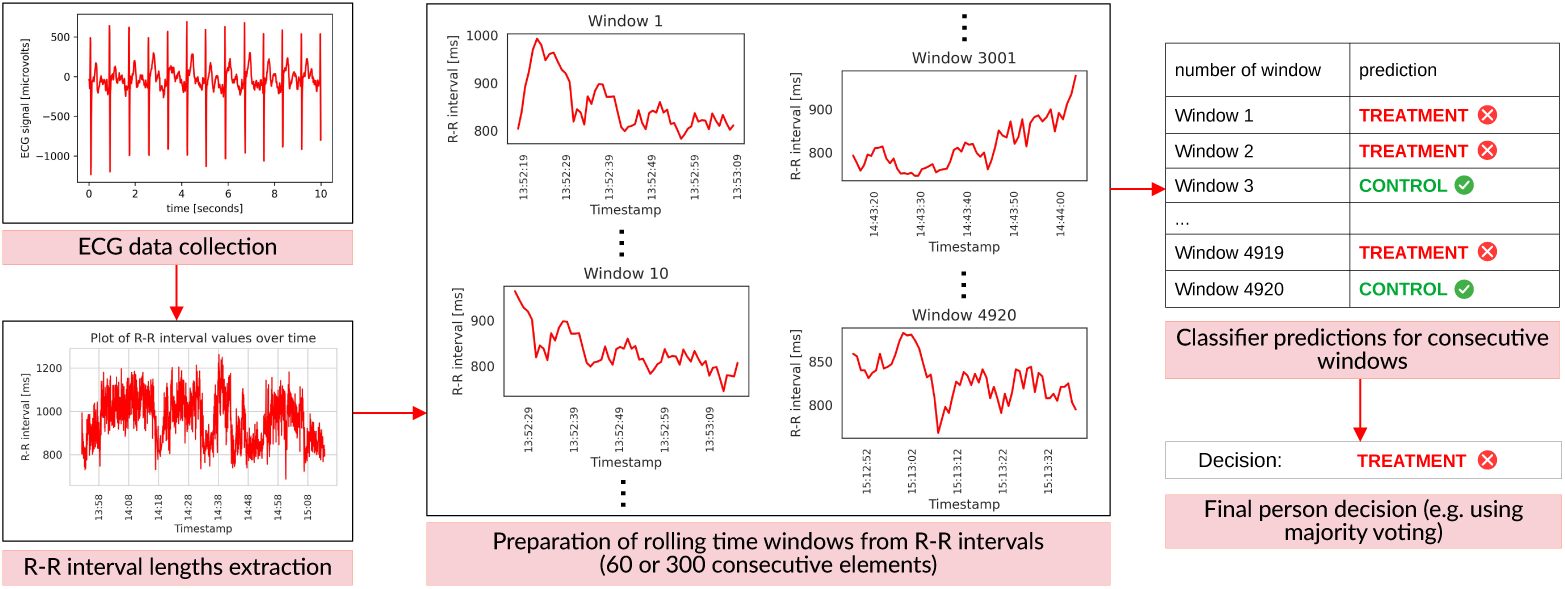
The scheme of the presented approach. First, for each person, ECG data is collected, from which a sequence of R-R interval lengths is extracted. Next, we apply a rolling window of size 60 or 300 to the aforementioned sequence of R-R interval lengths. The step between two consecutive windows is one, therefore there is an overlap of 59 or 299 R-R values between the two consecutive time series extracted by our rolling window algorithm. Then, for each window, a classifier predicts whether it belongs to the person from the control or treatment group. Finally, a single decision for a given person is returned, based on predictions for multiple windows.

## 2 Methods

### 2.1 Dataset

The dataset used for the classification experiments contains 1-1.5 hour recordings of R-R intervals for 60 participants, collected with a Polar H10 ECG chest strap suitable for measurements under different activity conditions [36]. Out of the 60 participants, 30 belong to the control and 30 to the treatment group. The age range is 20 to 69 years in the treatment group and 24 to 69 years in the control group. In the patient group, 23 have been diagnosed with schizophrenia and 7 with bipolar disorder. Qualified psychiatrists diagnosed all study participants according to ICD-11 guidelines. We decided to analyze schizophrenia and BD together as both are psychotic disorders with similarities in underlying mechanisms and HRV changes. For further explanation, please refer to Section 4.1. The sex structure in both groups is comparable, i.e. the treatment group consists of 16 women and 14 men, while the control group includes 17 women and 13 men. The dataset is publicly available in [51] while the experimental procedure was accepted the Bioethics Committee of the Medical University of Silesia, No.: BNW/NWN/0052/KB1/135/I/22/23.

Based on the analysis depicted in [39], all participants were examined using the PANSS questionnaire designed for the assessment of symptom severity in psychotic disorders [52]. The mean values of consecutive PANSS subscores equal 16.37, 19.80, 35.30 and 71.47 for positive, negative, general and total scores, respectively. Furthermore, as shown in Fig. 13 in [39], HRV was negatively correlated with the general score of PANSS (*r* = 0.45, *p* = 0.01). A more detailed evaluation of this dataset was performed in [39].

To perform an unbiased classifier evaluation, we divided the dataset into five separate folds in a standard cross-validation scheme, with windows containing a maximum of 60 or 300 consecutive R-R interval lengths, which correspond approximately to 1- or 5-minute recordings. Therefore, we have a total of 545604 or 521204 individual time-rolling windows with step length 1, respectively. Thus, there is an overlap of 59 or 299 R-R interval lengths between two consecutive time series.

During each of the five cross-validation steps, one fold is selected as a test set, while the remaining four folds are used for training and validation sets. A single fold contains all windows of six patients and six control individuals. This approach prevents data leakage between folds, as all measurements of a given individual are confined to a single fold. Training is performed in two phases: In the first phase, hyperparameters are optimized, while in the subsequent phase, a model is trained with the appropriate hyperparameter values. In the initial phase, during hyperparameter optimization, one of the four training folds is designated as the validation set. Once the optimal set of hyperparameters is determined, the four training folds are combined. During the final training phase, the data split into training and validation sets (within the four training folds) varies depending on the method, while the test set remains consistent across all methods.

For clarification, in Fig. 2, we depicted the distribution of individual R-R intervals’ lengths for the treatment and control groups for consecutive folds with 60-element time windows (consisting of approximately 1-minute long measurements), as well as median R-R values with the corresponding standard deviations.

**Fig 2.**
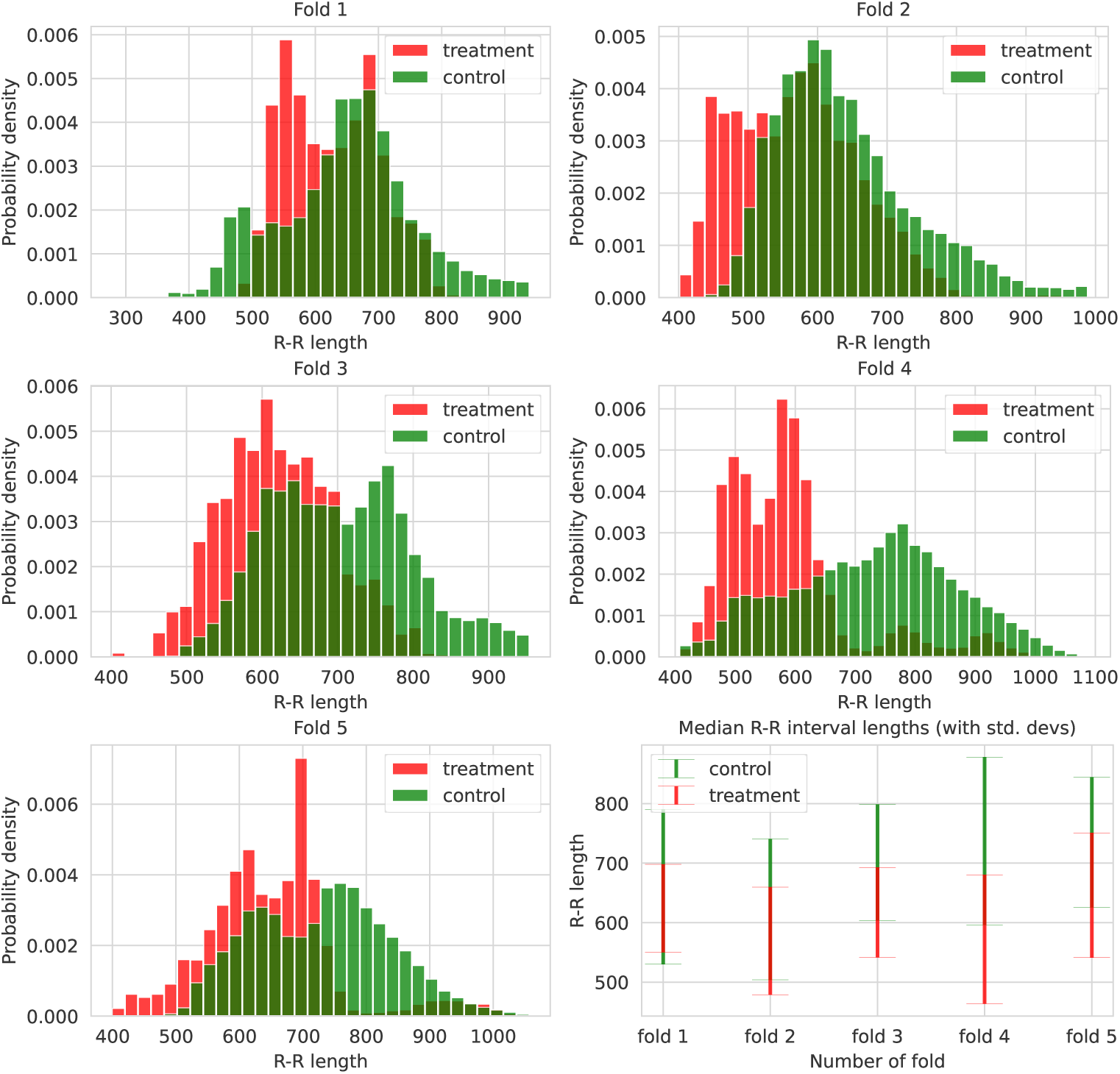
The distributions of 5 folds designed for the cross-validation classification experiments with 60-element time rolling windows. Each fold contains 6 patients and 6 persons from the control group, and when merged, they form the whole dataset. In consecutive iterations, samples (sequences of R-R interval lengths) from four folds form a training and a validation set, while the remaining fold constitutes a test set.

The presented plots indicate differences in the distributions between folds. Although folds 1, 3, and 5 are quite comparable, folds 2 and 4 are slightly different from them. For instance, fold 4 contains lower individual R-R values from the treatment group and higher control R-R values than the remaining folds. Such heterogeneous data are demanding for machine learning models, but the differences in data distribution are often inevitable in real-world cases with a limited amount of data. However, we did not want to manually modify the people selection in consecutive folds to avoid selection bias and ensure reliable and fair experiments. The assignment of persons to folds was performed completely at random.

Our approach handles a real-world scenario in which long-term monitoring of many patients is challenging because patient signals have various characteristics. Performing experiments without severe limitations for evaluated persons (e.g., being in an isolated environment) is easier to arrange in practice but harder to analyze further. However, 60 persons with 1-1.5 hour recordings deliver a large number of individual time windows. We anticipate that brief events that alter the lengths of the R-R intervals, such as short walks, as described in the experiment procedure in [39], will not prevent the classifier from making accurate diagnoses for a substantial number of windows.

We also performed an additional leave-one-out cross-validation experiment to verify the impact of a larger and more diverse training set on the test results. In this scenario, recordings from 59 people form the training set during each training run, and the remaining person is in the test set. The procedure is repeated 60 times, each time for a different test person. This experiment is considerably more time-consuming than the 5-fold cross-validation and is therefore used solely as a supplementary analysis.

### 2.2 Classifiers

From the theoretical point of view, our task, the diagnosis of schizophrenia based on the R-R interval lengths, can be seen as a time series classification task, for which various approaches have been introduced, including methods based on neural networks [53], Bayesian networks [54], hidden Markov models [55], genetic algorithms [56], Support Vector Machines [57], decision trees [58], frequent patterns (known as *motifs* or *shapelets*) and hubness-aware classifiers [59]. In our study, we considered the most prominent approaches that will be described in the following sections.

#### 2.2.1 Nearest neighbor

In [60], authors compared various time series classifiers, and concluded that 1-nearest neighbor “is an exceptionally competitive classifier”. For this reason, it has been widely considered a baseline in the last decades. Although the nearest neighbor classifier had been used mainly with dynamic time warping (DTW) distance, it was shown that the Euclidean distance may lead to similarly accurate results [61]. In particular, with increasing dataset size, the difference between the accuracy of the nearest neighbor with DTW and Euclidean distance is decreasing, and in the case of moderately large datasets, the difference is negligible [62]. At the same time, the Euclidean distance can be calculated in orders of magnitude faster than DTW, especially if the calculations are performed on GPUs.

For these reasons, we considered 1-nearest neighbor with Euclidean distance as a baseline in our experiments. For computational efficiency, we randomly selected 100000 training time series and searched for the nearest neighbors of test/validation time series only among those selected time series. Additionally, in line with the experiments with other models, this allowed us to repeat the experiments with various seeds that led to various selections of the training time series. The only hyperparameter we learned using the validation data was the decision threshold for classifying persons, i.e., the ratio of positively classified segments, so that the person will be classified as positive.

#### 2.2.2 Multilayer Percepton

Multilayer perceptrons (MLP), i.e. fully connected feed-forward neural networks, with at least one hidden layer and non-linear activation, are known to be universal function approximators [63]. Therefore, MLP served as a deep learning baseline in our experiments. In particular, we considered MLP with a single hidden layer containing 128 units. We used the sigmoid activation function in the hidden layer and softmax activation in the output layer. We trained the model for 100 epochs using the Adam optimizer with cross-entropy loss. We learned the decision threshold for window classification, the decision threshold for person classification, as well as the most important hyperparameters of the optimizer, i.e., the learning rate and batch size, using the validation data. In particular, in each round of cross-validation, we performed a grid search and we searched for the best thresholds from {0.01, 0.02*, …,* 0.99}, learning rate from {10*^−^*^3^, 10*^−^*^4^, 10*^−^*^5^}, and batch size from {256, 512, 1024}.

#### 2.2.3 SVM + FSH (feature selection with automatically configured hypothesis tests)

The method follows a classical approach to feature engineering with feature extraction and selection based on hypothesis tests. Initially, feature extraction is conducted using the “efficient” set of *tsfresh* [64] features, comprising over 700 features. Subsequent feature selection involves eliminating features with low variance and statistically identifying the 50 most pertinent features via the Mann-Whitney U test. Preprocessing is performed through robust normalization and random subsampling, in which the classifier is trained on a balanced subset of 1500 examples from each class. The classification task is executed utilizing an SVM with an RBF kernel. Parameter optimization for the SVM’s C and gamma hyperparameters is achieved through a grid search in the range from 10*^−^*^1^ to 10^2^, based on the model’s performance on the validation set.

The threshold estimation for person classification is approached heuristically, determining it as the midpoint of the median scores across the classes derived from the training and validation sets.

#### 2.2.4 Ensemble of SVMs

The ensemble classification scheme for R-R interval data follows the bagging paradigm utilizing a set of SVM classifiers 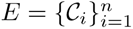, each trained on a different and exclusive subset of training data with hyperparameters *C, γ* determined by the method described in [65]. Those classifiers then perform a majority vote on the classification of each window in the test set.

Classification of a person is obtained by determining whether a window classified as class “positive” exceeds the threshold determined on the validation set in the following way. For each person in the validation set, a *R_i_* percentage of windows classified as ‘*i*’ is calculated, where *i* ∈ {0, 1}. Thus, *R_i_*’s are cleared of outliers, understood as data more distant from the mean than 2 standard deviations. Finally, the threshold *t* is set as the mean between the average values in *R*_0_ and *R*_1_.

Data preprocessing is performed for each window *w* by extracting features: min(*w*), max(*w*), var(*w*), mean(*w*), median(*w*).

#### 2.2.5 XGBoost

XGBoost [66] or Extreme Gradient Boosting, is a scalable and efficient implementation of gradient boosting that has proven highly effective in various machine learning competitions and tasks. XGBoost like the other tree-based methods is very prone to overfitting, thus initially we used grid search to determine the hyperparameters. Two hyperparameters were checked, *NEstimators* and *MaxDepth*. The *NEstimators* parameter specifies the number of gradient-boosted trees used in the model, directly influencing the complexity of the model and its potential to overfit or underfit the data. The *MaxDepth* parameter controls the maximum depth of the trees, affecting the model’s ability to capture complex patterns and its risk of overfitting with deeper trees. Using full cross-validation for each set of hyperparameters we trained the model on *NEstimators* ∈ {10, 50, 100, 200, 500} and *MaxDepth* ∈ {5, 10, 15, 20, 25, 30, 35, 40, 45, 50}. Then, we used the accuracy of the validation set to select the optimal hyperparameters. The optimal hyperparameters were *NEstimators* = 200 and *MaxDepth* = 5 in both cases, i.e. for 60 and 300-element time series. In the second stage, we used the validation subsets to determine the percentage of “positive” predictions that correspond to the highest person accuracy. If more than one value corresponded to the highest accuracy, we chose the value closest to 50%. The determined values were 49% and 44%, for 60 and 300-element windows, respectively.

#### 2.2.6 GRU + FCN

Another deep learning architecture selected for the classification experiments was GRU+FCN, i.e. Deep Gated Recurrent and Convolutional Network Hybrid Model [67], designed for univariate time series classification. The model architecture consists of two independent paths, the first of which is created by GRU blocks followed by the dropout layer, while the second part is formed by three blocks of convolutional layers followed by batch normalization and the ReLU activation in each of the CNN blocks. At the end of the second path, a global average pooling is applied. Finally, the output of the two model parts is merged and a fully connected layer assigns a label to a given sample. The authors of GRU+FCN proposed its approach based on LSTM+FCN [68], replacing LSTMs with GRU cells. We decided to include a recurrent neural network architecture in our studies because such an approach is well-suited for time series analysis, capturing short- and long-range dependencies.

For both 60- and 300-second time windows, we trained models through a maximum of 30 epochs with early stopping (based on the validation loss value), 10 epochs of patience and a learning rate scheduler with 5 epochs of patience. After that, the learning rate was reduced by an order of magnitude. We performed optimization of the crucial hyperparameters using all five folds of the dataset, whose preparation has been described in Section 2.1. We searched for an initial learning rate from the set {10*^−^*^4^, 10*^−^*^3^}, batch size among {64, 128}, the number of GRU layers {1, 2}, the hidden state sizes {100, 200}, dropout probability (in the GRU path) {0.4, 0.8}, as well as for uni- and bidirectional recurrent networks. Furthermore, for the 60-element time windows, a batch size of 16 was evaluated.

As the highest-performing setup, we selected a network having a single layer of the unidirectional gated recurrent unit followed by a dropout layer with a probability of zeroing out values set to 0.8. Also, the convolutional layers were adjusted to 128, 256 and 128 kernels. The kernel sizes were set to 7, 5 and 3, respectively. For 60-element time windows, we selected an initial learning rate of 10*^−^*^4^, a batch size of 128 and a hidden state size in GRU equal to 100. However, for the larger, 300-element time windows, an initial learning rate was about an order of magnitude higher than previously, that is, 10*^−^*^3^, a batch size was reduced to 64 and a hidden state of GRU was extended to 200.

## 3 Results

### 3.1 Cross-validation experiments

In Table 1, we present results for all the evaluated machine learning methods in the case of 5-fold cross-validation experiments. As evaluation metrics, we selected window and person accuracies. More specifically, we compared the accuracy of all consecutive time windows with the accuracy for a given person based on a criterion specific to the method, e.g. majority voting. It is necessary to emphasize that not all windows for an evaluated person must be correctly classified to determine whether such an individual belongs to the control or treatment group.

**Table 1.**
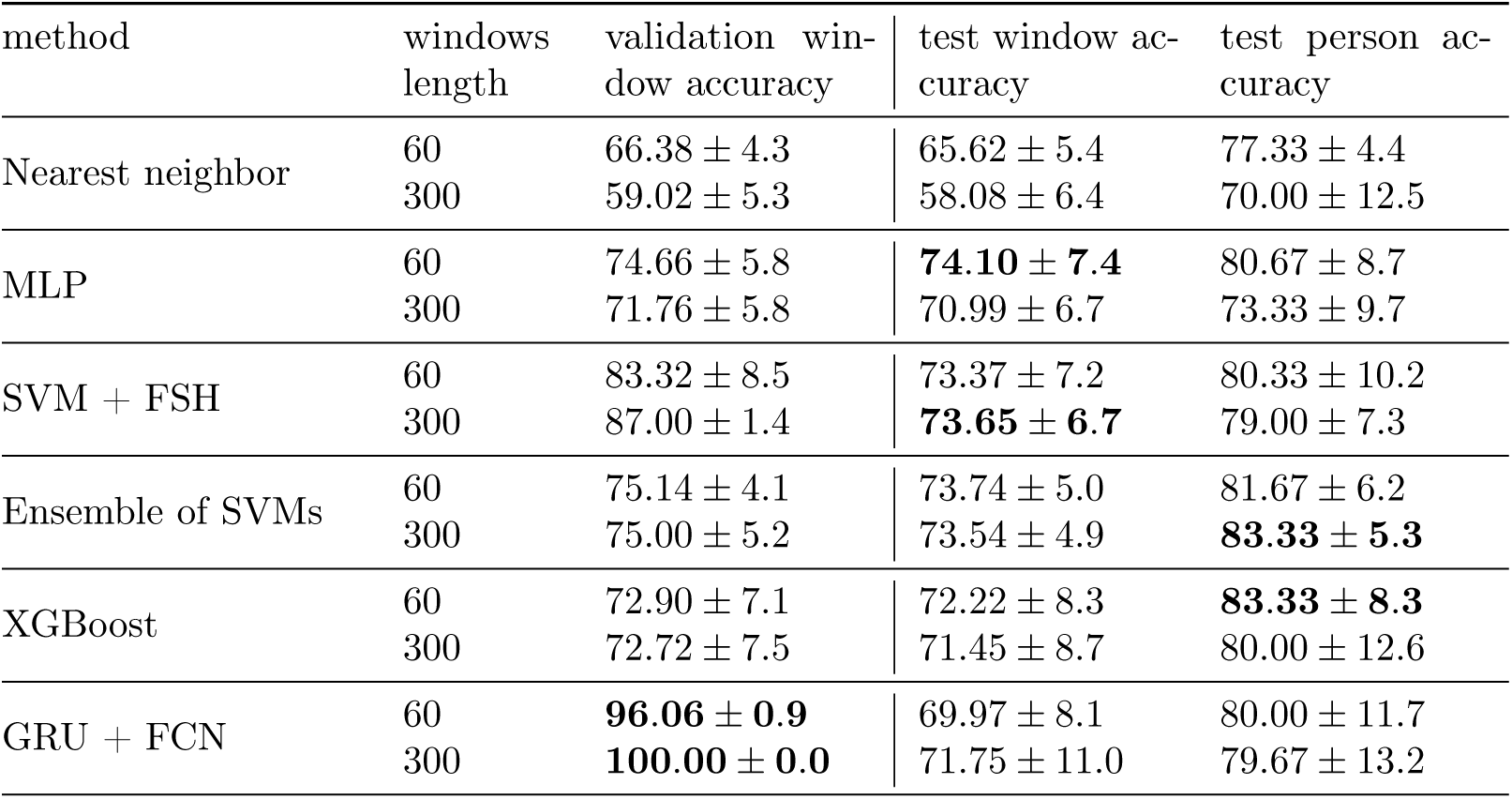
Experimental results for different methods and time window lengths averaged over 5 test folds, in terms of overall accuracy [%]. Also, in the case of the non-deterministic methods, results are additionally averaged over 5 training runs per each cross-validation step. Bold font indicates methods with the highest accuracy in a given category. MLP − multilayer perception; FSH − feature selection with automatically configured hypothesis tests; GRU − Gated Recurrent Unit; FCN − fully convolutional network.

We compared two variants of the time window length, i.e. 60 and 300 consecutive R-R interval lengths in a single window. Analysis reveals that the two considered time window length variants yield comparable performance, suggesting that providing additional contextual information, such as extending measurements to approximately five minutes, is unnecessary for the classifiers. This conclusion is important from a practical point of view because 300-element time windows are more computationally expensive while not bringing clear advantages. Also, most of the methods perform even slightly better for shorter windows.

We also noticed that the methods based on the ensemble of classifiers like XGBoost and Ensemble of SVMs lead to the highest test person accuracy, exceeding 80% for both compared window settings. Taking into account that the final person’s decision may be based on multiple time windows, this experimental scenario is more favorable for ensemble methods. They are designed to benefit from the variety of information contained in different windows. Their performance is the highest, even though the accuracy for individual windows is slightly higher for MLP and SVM + FSH. However, it is necessary to emphasize that the differences between the compared methods are not huge and, in general, they achieve similar accuracy, except for the nearest neighbor classifier having the lowest scores in each category.

It is important to highlight the significant disparity between validation and test window accuracies for the GRU + FCN model. This approach led to the best results for validation windows, achieving even 100% for 300-element windows, but it has no real advantage during the evaluation on test windows. It suggests that the discrepancy between the validation and test folds hinders the full potential of these methods from being realized, which was discussed in Section 2.1.

Nevertheless, in such an experimental setting, which corresponds to the real-world scenario with limited possibilities of patient monitoring, we can still classify 8 out of 10 evaluated persons correctly, on average.

We performed an additional series of experiments in a leave-one-out cross-validation scenario for two methods, i.e. Ensemble of SVMs and GRU + FCN. Their results are presented in Table 2 while the distributions of consecutive in-person accuracies for 60 test folds are depicted in Fig. 3. The two analyzed methods achieved comparable performance, both in terms of test window accuracy (70.1% for Ensemble of SVMs and 71.8% for GRU + FCN) as well as for test person accuracy (80% for Ensemble of SVMs and 79.4% for GRU+FCN). As with previous results, the GRU + FCN model achieved exceptionally high accuracy in the validation windows and demonstrated near-perfect performance. However, its accuracy declined on the test windows. Based on the presented histograms we can observe that for the majority of persons, more than 50% of windows were classified correctly. Also, the median fold accuracy was equal to 76.3% for GRU+FCN and 77.9% for Ensemble of SVMs.

**Fig 3.**
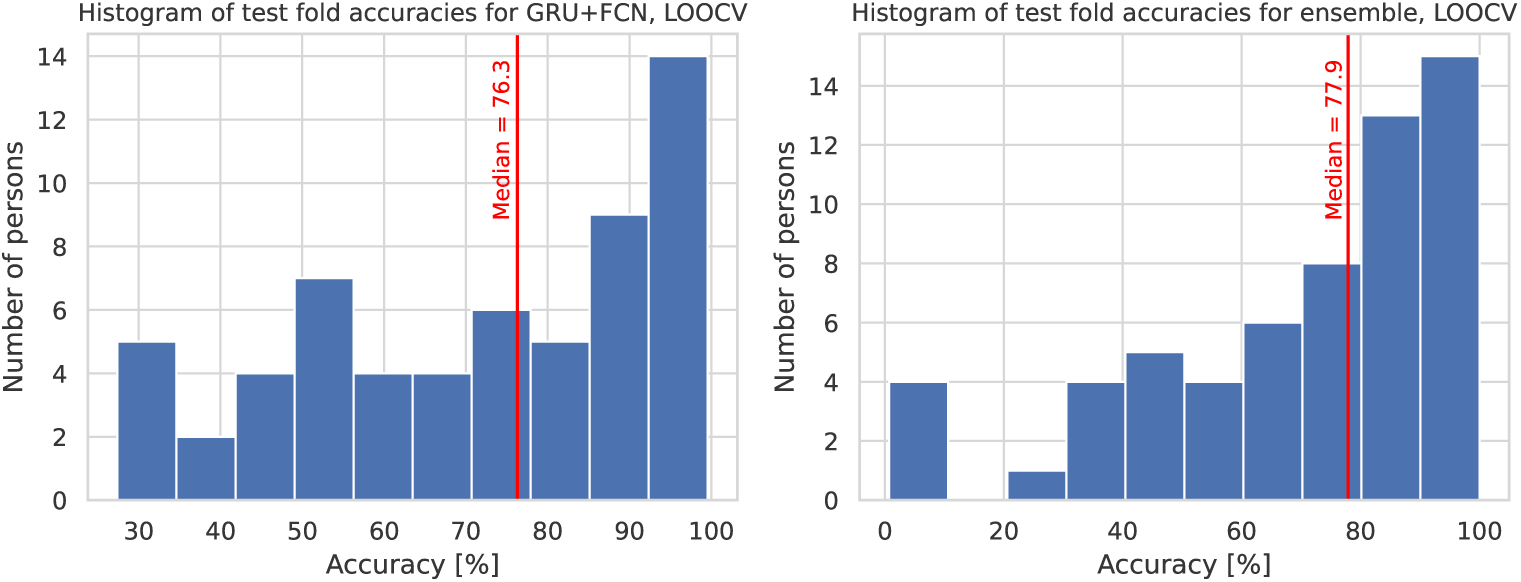
The distributions of 60 test fold accuracies for the leave-one-out cross-validation experiment for GRU+FCN (averaged over three different runs) and Ensemble of SVMs methods.

**Table 2.**
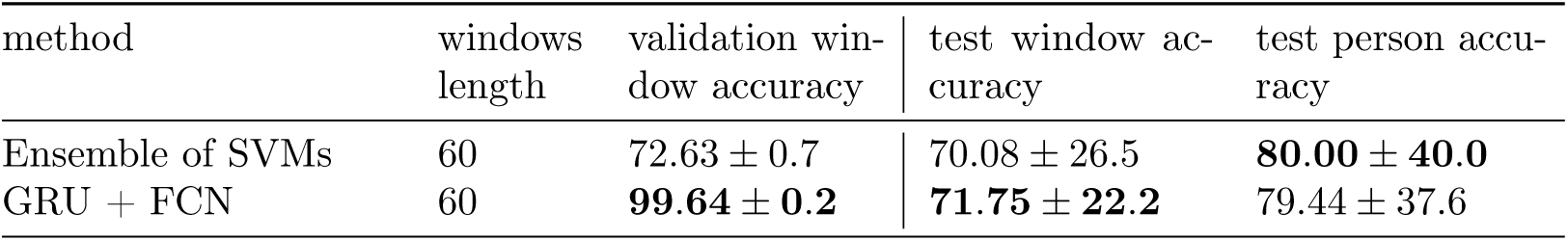
The results of the leave-one-out cross-validation experiment averaged over 60 test folds (in terms of overall accuracy [%]). In the case of the non-deterministic method, like GRU + FCN, results are additionally averaged over 3 training runs per each cross-validation step. Bold font indicates the highest-performing method in each category.

| method | windows length | validation window accuracy | test window accuracy | test person accuracy |
| --- | --- | --- | --- | --- |
| Ensemble of SVMs | 60 | $72.63 \pm 0.7$ | $70.08 \pm 26.5$ | **$80.00 \pm 40.0$** |
| GRU + FCN | 60 | **$99.64 \pm 0.2$** | **$71.75 \pm 22.2$** | $79.44 \pm 37.6$ |

### 3.2 The impact of data features on classifier decision

To analyze the influence of features on model decisions, we employed SHapley Additive exPlanations [69] (SHAP). SHAP quantifies each feature’s contribution to a model’s prediction (here, the probability of belonging to the positive class), ensuring that, for every instance, the sum of SHAP values for all features equals the difference between the model’s output and its baseline (average) prediction.

Although the models presented in this article use the raw data and learn the features themselves, for this investigation, we have manually selected a small set of seven distinct, well-known and intuitive features: mean, minimum, maximum values, standard deviation, skewness, kurtosis, lag-*i* autocorrelation (ACF) values in each window, where *i* ∈ {1, 2, 3, 4}. We then applied a random forest (RF) classifier to a dataset resampled by selecting every 500th instance (so the visualization is not overloaded). In a standard 5-fold cross-validation experiment performed on this subsampled dataset, the RF classifier achieved 73.3% accuracy with 60-element windows and 73.9% with 300-element windows, suggesting that these features are effective data descriptors. The importance of features obtained was similar in both experiments, so only the 60-element windows’ experiments are presented here.

Fig. 4 presents the bee-swarm plot that highlights the distribution of feature impact on model predictions and depicts how this impact correlates with feature values (high/low). It reveals that standard deviation and maximum values are the most essential features, with high values (red) linked to the negative class (control group), and low values (blue) related to the positive class (treatment group), especially for standard deviation. This tendency agrees with our observations that the low variance of R-R values and shorter beat-to-beat intervals are characteristic of the treatment group.

**Fig 4.**
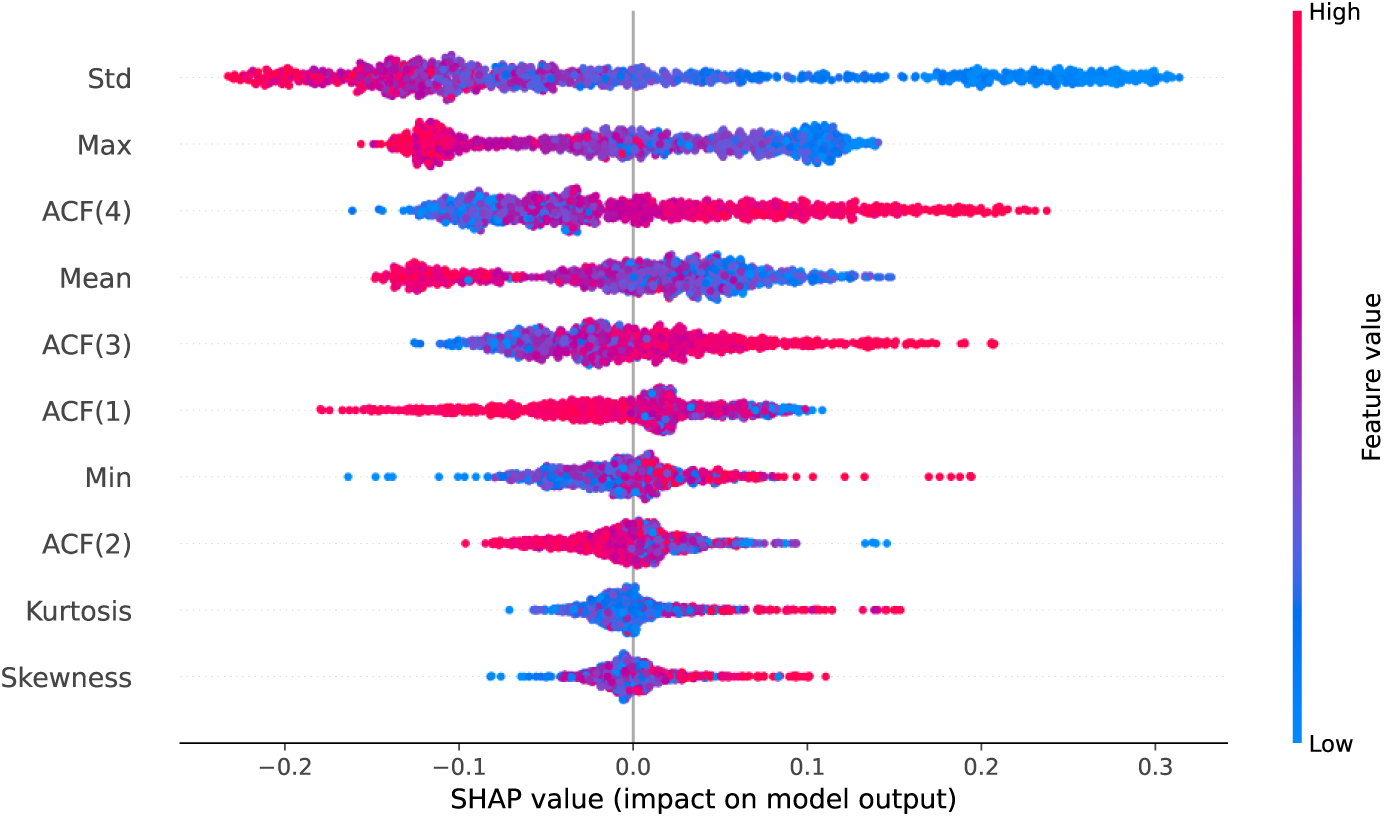
The distribution of feature impact on model predictions of the treatment (positive SHAP values) and the control group (negative SHAP values) as a SHAP bee-swarm plot. Every point represents a data instance with clusters indicating the data density. Rows and columns correspond to features and SHAP value, respectively, while color intensity represents the feature value. The red color indicates high values of the considered features while the blue color represents the opposite case. Features are ranked by their impact on the model output. Data is from a 60-element windows’ experiment, on a dataset sampled every 500th instance.

Lag-*i* ACF values, where *i* ∈ {1, 2, 3, 4}, show relationships of the currently considered R-R interval to their first, second, third and fourth neighboring R-R interval values, respectively. These variables are considerably important in terms of influencing the model predictions. High lag-4 and lag-3 ACF values are associated with the positive (treatment) group, while high lag-1 and lag-2 ACF values are related to the control group.

We extended our analysis concerning ACF and separately calculated its values for all data instances, as presented in Table 3. The highest autocorrelation is observed between two consecutive R-R intervals. However, its mean values are higher for the control than for the treatment group, especially for the 60-element time series (0.84 ± 0.1 vs 0.78 ± 0.2). This tendency reverses for larger lag values, and finally, for lag-4, ACF is much higher for the treatment group (0.74 ± 0.17 vs 0.61 ± 0.21 for 300-element time series) than for controls. These observations are further discussed in Section 4.3.

**Table 3.** Autocorrelation function with lag values, i.e. ACF(*i*), where *i* ∈ {1, 2, 3, 4}, for control and treatment groups across all time windows. For lags greater than 2, the mean ACF values for the treatment group consistently exceed those of the control group.

| lag | window | mean ± standard deviation | |
| --- | --- | --- | --- |
|  |  | control group | treatment group |
| ACF(1) | 60 | **0.84** ± 0.12 | 0.78 ± 0.20 |
| ACF(1) | 300 | **0.91** ± 0.07 | 0.90 ± 0.11 |
| ACF(2) | 60 | **0.67** ± 0.19 | 0.65 ± 0.25 |
| ACF(2) | 300 | 0.81 ± 0.12 | **0.84** ± 0.13 |
| ACF(3) | 60 | 0.51 ± 0.23 | **0.58** ± 0.24 |
| ACF(3) | 300 | 0.70 ± 0.17 | **0.79** ± 0.15 |
| ACF(4) | 60 | 0.37 ± 0.27 | **0.50** ± 0.24 |
| ACF(4) | 300 | 0.61 ± 0.21 | **0.74** ± 0.17 |

Kurtosis and skewness have a visibly lower impact on the classifier decision, which may result from the fact that R-R intervals, especially within a short window, are likely to have a relatively symmetrical distribution if the heart rate does not change significantly due to external factors.

Fig. 5 presents the heatmap plot where the X-axis corresponds to the instances ordered by the model output value (sum of SHAP values) shown at the top of the plot. We can see how the examples for which the model is most confident in its decision are mainly impacted by a small subset of features and how other features are more relevant for less obvious instances located in the middle of the presented X-axis range.

**Fig 5.**
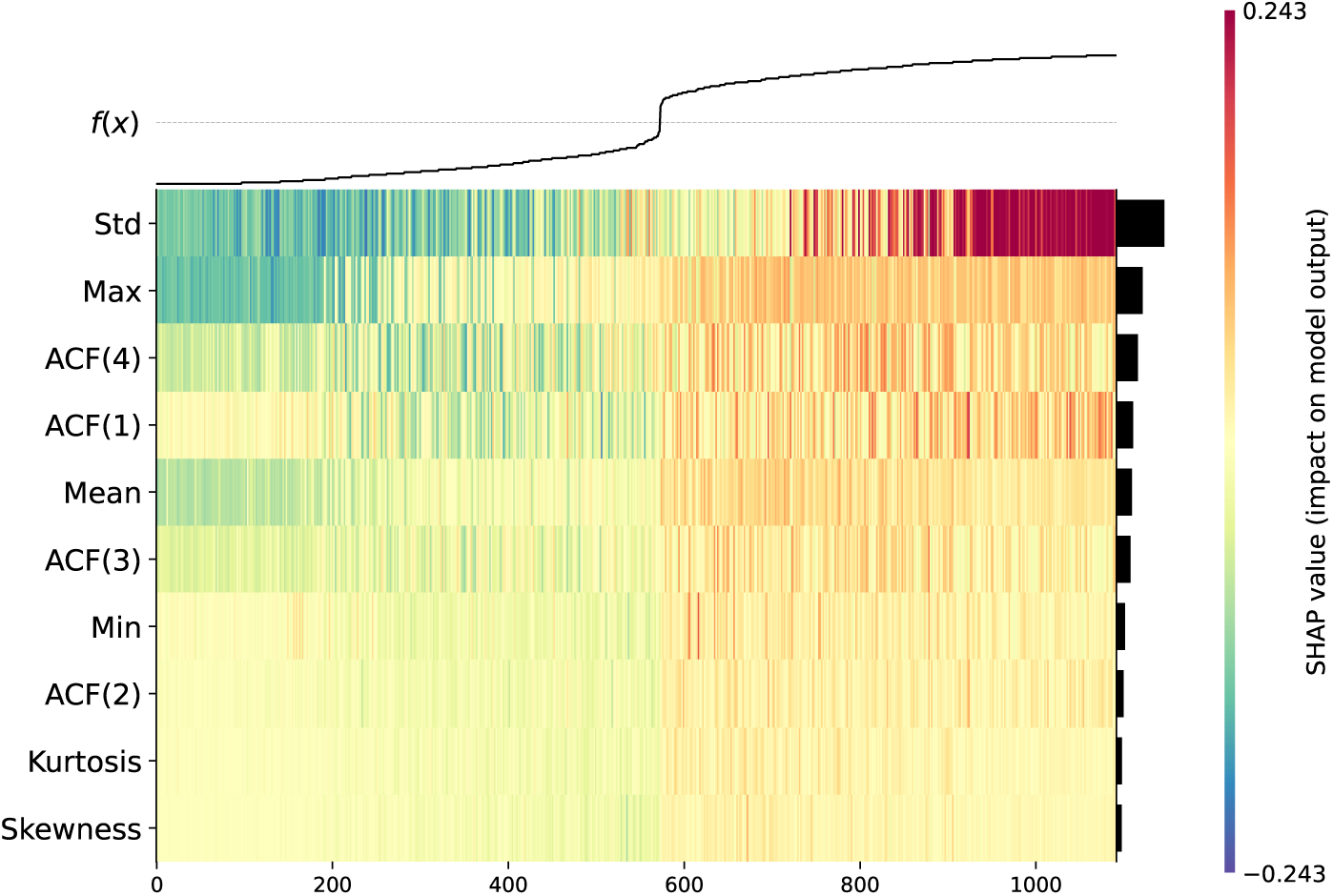
The influence of features on model predictions, presented as a SHAP heatmap. Rows and columns correspond to features and instances, respectively. Instances are sorted according to the model’s output score. Color intensity represents the SHAP value, with red indicating a positive impact and green a negative impact on the model output (probability of belonging to the positive class) relative to the baseline. Features are ranked by their importance and are also presented as sidebars. Data is from a 60-element windows’ experiment, on a dataset sampled every 500th instance.

### 3.3 Classifier diversity and ensemble methods

Due to the diverse patterns among evaluated individuals, each scenario contains a subset of data that is misleading for classifiers. Although the evaluated methods are consistent in straightforward cases, as shown in the first plot in Fig. 6, they generate numerous errors in ambiguous cases, resulting in a high number of misclassified time windows like for the individual from the middle plot in Fig. 6). Classifiers occasionally disagree, as presented for the bottom individual, for which GRU+FCN incorrectly classified most windows, whereas Ensemble of SVMs was correct in nearly all its predictions.

**Fig 6.**
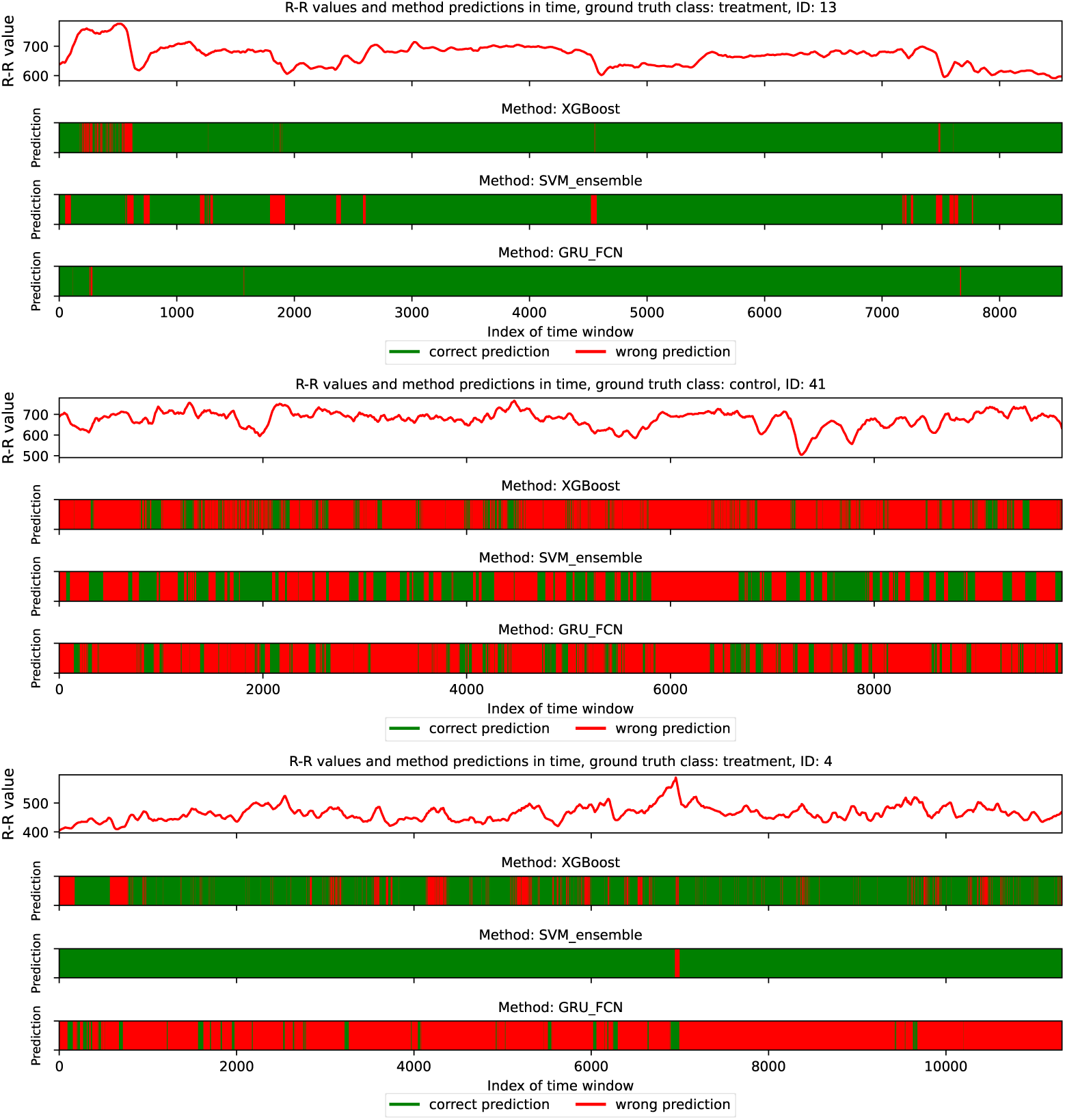
The overview of R-R values and predictions of XGBoost, Ensemble of SVMs, and GRU + FCN classifiers for consecutive time windows for the three selected individuals. Green areas correspond to the correctly classified periods while red areas refer to the opposite case. The first plot depicts the individual from the treatment group being relatively straightforward for all three classifiers. The middle plot corresponds to the selected control group individual for which most of the time windows are classified incorrectly by all three compared methods. Finally, the last plot represents the treatment group individual classified mostly accurately by XGBoost and Ensemble of SVMs but misclassified by GRU+FCN.

This observation aligns with the no-free-lunch theorem, which states that there is no universal classifier with high performance for diverse datasets. Although each evaluated method may focus on different patterns, even for correctly classified individuals, a subset of time windows is misclassified due to locally varying characteristics. This considers patients in both movement and rest conditions. For example, for control individuals, the R-R intervals decrease during activity and exhibit low variance within the analyzed windows, making them more similar to the individuals in the treatment group, who generally have shorter R-R interval lengths with slight variance (see Fig. 5 and the upper plot in Fig. 7).

**Fig 7.**
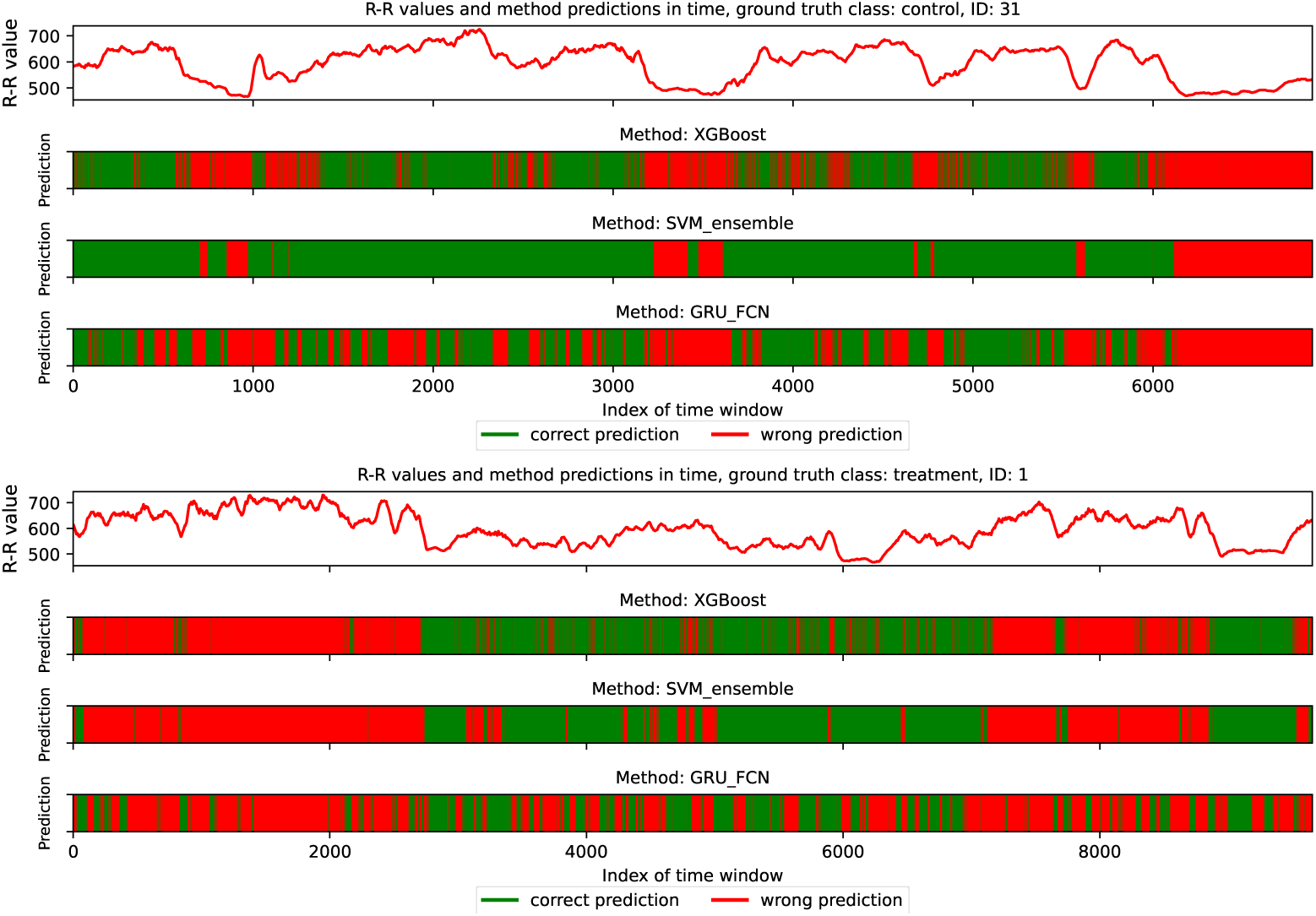
The overview of R-R values and predictions of XGBoost, Ensemble of SVMs, and GRU + FCN classifiers for consecutive time windows for the two individuals. Green areas correspond to the correctly classified periods while red areas refer to the opposite case. The upper plot corresponds to the control group individual for whom selected contiguous time windows were classified incorrectly. All three tested methods made errors for periods corresponding to lower R-R interval lengths, while for the remaining time windows, the Ensemble of SVMs achieved the highest performance. The lower plot depicts the individual from the treatment group whose signal is demanding for classifiers. Only the middle part of the measurements mostly led to the correct labelling (except for GRU + FCN making many prediction errors but still less than for the remaining parts of the signal).

#### Stacking classifier

The high level of disagreement between classifiers (see e.g. the bottom plot in Fig. 6) raises the question of whether forming a stack of classifiers would improve classification accuracy. To investigate this, we conducted an additional experiment using a stack of three methods: GRU+FCN, Ensemble of SVMs, and XGBoost, with a logistic regression classifier. The classifier input to the logistic regression model was a three-element sequence of 0’s or 1’s based on the predictions of each method for a given time window. However, the final classification accuracy remained similar to the best result among the individual models, indicating that, in this case, the use of multiple methods does not significantly improve overall results.

#### Long-term predictions

The time windows in this study were approximately 1 or 5 minutes in duration, corresponding to 60 or 300 consecutive R-R intervals, respectively. Short-term measurements in randomly selected time windows may lead to incorrect predictions in changing conditions. However, in many cases, classification with 60-element time windows yielded higher accuracy than for the 300-element sequences, as presented in Table 1. Therefore, extending the time range within individual windows seems to be unnecessary. Nevertheless, as shown in Fig. 7, some time periods may be easier to classify than others. Thus, the selection of windows merits consideration in future experiments and may increase the classification accuracy.

#### Minimum variance critertion

We evaluated an alternative voting strategy (compared to the one described in Section 2.2) for assigning the final label which relies on identifying the most extended period of stable predictions characterized by the lowest prediction variance. This approach is based on the premise that consistent predictions over a long time period should provide more reliable information than predictions across varying states. To evaluate this, the variance was calculated based on 60 consecutive time window predictions. Then, the longest time range having the lowest variance was considered, and the most common class within this subset was set as a model prediction.

The experiment was carried out using the three methods described in Section 2.2: Ensemble of SVMs, XGBoost, and GRU + FCN. This strategy improved the accuracy of XGBoost to 85.00 ± 12.25%, the highest result among all methods tested. However, as it did not improve the performance of the other methods, the approach was ultimately not adopted.

## 4 Discussion

### 4.1 Schizophrenia and bipolar disorder as a common group of psychotic disorders

Although schizophrenia and bipolar disorder are classified as two distinct diseases based on the phenomenological description of symptoms [9, 10], there is an ongoing debate whether both diseases actually represent a continuum [70, 71]. Based on MRI findings on the neurological level, both diseases share a common altered network of brain areas [70], both manifest as psychotic disorders [37] and both share many common genetic, endophenotypic and phenomenological traits [38]. One of such prominent endophenotypic traits is the shared difference in ANS regulation in comparison to healthy controls as observed through heart rate variability that is well described in both schizophrenia [21] and BD [72]. For these reasons, we chose to analyze both diseases collectively, aligning with other studies that examine these groups together with respect to severity of symptoms [32] or by broadly defining the studied population as psychotic [73].

### 4.2 The effect of psychotropic medications on classification results

The influence of antipsychotic medications on HRV is well established, with a diminishing ANS activity and a decrease in HRV, especially in the patient group taking quetiapine [74]. This raises the question of whether classifier results actually represent a distinction between psychotic patients and healthy controls, or rather between patients under antipsychotic medication and healthy controls. Due to bioethical concerns, it is not possible to withdraw the medication during hospitalization. However, medication is not the only or even the main reason for HRV changes in the patient group. Firstly, there are differences between the influence of different antipsychotic drugs on ANS [39, 74]. Patients in our group were treated with a range of antipsychotic medications. Given that different antipsychotics exhibit distinct pharmacological profiles and varying effects on the autonomic nervous system, the treatment group is likely to exhibit greater heterogeneity, potentially complicating classification efforts. However, it is well established that psychotic patients have changes in ANS activity and HRV [21, 72]. In addition, a recent work considering this dataset [39] indicated that HRV is related to the severity of psychotic symptoms. Even if antipsychotic medications influence the features utilized by the classifiers, this phenomenon does not undermine the significance of ongoing research efforts in this area. In such cases, these classifiers would help clinicians distinguish between patients who comply with treatment guidelines and those who have stopped taking their medicine and are at risk of disease remission.

### 4.3 Relationship between ACF values and ANS balance

Based on the SHAP values presented in Fig. 4, we can observe that the high lag-3 and lag-4 R-R interval autocorrelation values are more common in the treatment group, while the high lag-1 and lag-2 autocorrelation are mostly related to the control group. This may be partially explained by the fact that parasympathetic and sympathetic regulation of HRV through the ANS occurs at different time intervals, with a maximum response of about 0.5 seconds for the parasympathetic branch and about 4 seconds for the sympathetic branch [75, 76]. Given that the autocorrelation functions (ACFs) of successive lags correspond to the aforementioned time intervals, these features have the potential to distinguish between parasympathetic and sympathetic activity. This is in line with the fact that the beat-to-beat regulation of HRV represented by lag-1 is parasympathetic in its nature [77] and that psychotic patients have an impaired parasympathetic response [21, 78, 79]. It suggests that lag-1 remains an important feature for the control group due to the proper functioning of the parasympathetic system and beat-to-beat regulation. Consequently, parasympathetic dysregulation would shift the balance in favor of a stronger sympathetic response in the patient group [21, 80], which could correspond to what we can observe as a growing strong relationship between the high value of the lag-3 and the subsequent lag-4 feature and the prediction in the patient group. In this case, the lower value of the lag-2 could represent the moment of transition between the peak parasympathetic and sympathetic response, in which neither the sympathetic or parasympathetic branch of the ANS has a dominant impact on this characteristic. This hypothesis is strongly supported by changes in the distribution of the impact of the feature on model predictions between subsequent lag ACF values.

There is an ongoing debate on whether the ANS imbalance in favor of higher sympathetic activity in psychotic patients occurs mainly because they have higher baseline sympathetic activity [80] or mainly because they have lower parasympathetic activity [21]. As we mentioned earlier, the parasympathetic system has a short response latency of 0.5 seconds and a return to baseline within approximately 1 second [76]. Therefore, the influence of parasympathetic activity on lag-1 and possibly also lag-2 should be high, with probably little to no impact on lag-3 and even less on lag-4. In the case of our results, given that the impact of lag-3 and lag-4 ACF on the prediction of the model is greater than that of lag-1 and lag-2 ACF, this could suggest that psychotic patients not only have a sympathetic shift in ANS balance but mainly have a higher baseline level of sympathetic activity compared to the control group. In a scenario where psychotic patients predominantly exhibit reduced parasympathetic activity, the influence of sympathetic activity, as reflected in lag-3 and lag-4 ACF values, would be expected to have a diminished impact on prediction within the patient group. Conversely, in the control group, the predictive contribution of lag-1 ACF, which represents parasympathetic activity, would be expected to be comparatively higher. However, this is not the case in our sample.

### 4.4 Relation of our study to other works

In this work, we have evaluated a diverse set of classifiers to generalize on particular characteristics of different algorithms. Additionally, our cross-validation scheme removes the bias of manual choice of hyperparameters. Our experiments address a broad spectrum of key methods in ECG processing, as outlined in [48]. Together, we view this as a solid foundation for quantification of the possible accuracy of a wearable HRV diagnostic system. The different methodologies employed by the authors of [41] and [42]complicate direct comparisons. However, our findings are broadly consistent with theirs and significantly enhance the generalizability of the evaluation of this diagnostic framework. Our paper supplements such works as [33] or [43], where no actual classification takes place.

When measuring HRV, an important variable is the length of the recorded sequence and patient activity. In this work, we use the dataset described in [39] which comprises relatively short time measurements (on the order of 1h) and lightly structured activity (patient-defined activity punctuated by walking). This limitation of time and broadly defined instructions are advantageous and provide flexibility (e.g. applicable to both on-site and at-home diagnostic) and provide minimal burden for the patient. We view this as a more accessible and versatile approach than long-term recording on the order of days [33, 41] or structuring the activity [42].

The diversity of patients is an important factor to consider when evaluating a diagnostic scheme. The dataset we considered is significantly larger (60 persons) than in previous works reporting classification results (24 in [41] or 28 in [42]), while simultaneously maintaining similar or higher accuracy in the case of GRU + FCN methods.

Our choice of sampling window (300 samples) corresponds to the theoretical analysis of short-term cardiorespiratory dynamics presented in [81], where it was found to provide a feasible assessment of cardiac dynamics variability from electrocardiographic records. Our experimental scheme is related to the one presented in [82], where three subsequent stress interventions were used, preceded by baseline recording and each followed by a rest period. In our case, walking interventions were used (see [39] for details), to achieve HRV tracking in a variety of activities.

### 4.5 Comparison of EEG- and ECG-based methods in psychiatric diagnostics

Most available research in the field of EEG in schizophrenia and BD focuses on the analysis of the electrical activity of the brain cortex in the resting state [83]. Consequently, machine and deep learning classification research in this field focuses mainly on resting state data [84, 85], partially because some of these EEG datasets are publicly available [86]. Another branch of EEG research in schizophrenia and BD is the analysis of event-related potentials (ERP), which are short EEG segments that are linked to particular events of experimental interest and are repeated through numerous trails to filter out signal noise [87]. One such ERP of particular interest is mismatch negativity (MMN), which has a well-documented link to psychotic disorders [88, 89]. Another ERP of potential use in psychiatry is the P300 component of auditory evoked potential, with some research showing its relationship to mood disorders, including BD [90]. ERP has potential as a tool for disease treatment and progression monitoring, mainly in the area of patient cognition evaluation [91].

DL-assisted whole brain resting state EEG activity has a very high diagnostic potential, although limitations come from its complexity and reluctance to adapt to clinical use by caregivers [84]. ML and DL methods using EEG tend to have a higher accuracy rate than ECG methods at the level of 90 to 99%, with few works reporting an accuracy rate of 70% [16]. Many of them are based on public sets, and even in the case of using one public dataset the accuracy of the methods between different studies may vary between 70 and 99% [16]. This may be the result of the richness of the data used in the studies. Also, it is important to remember that due to the complex spatial-temporal data structure, EEG research can often suffer from non-reproducibility due to inadequate data analysis methods and overfitting [92].

HRV focuses on changes in the beat-to-beat interval which are ANS activity-dependent [93, 94], while ECG signal morphology should be understood as the shape of the voltage curve over time. The morphology of the ECG represents different phases of depolarization and repolarization of the heart’s sinuses and ventricles. The authors in [95] analyze together different psychiatric disorders, including schizophrenia, and present various degrees of classification accuracy from 73.7 to 89.2%, based on the selected ECG lead, and accuracy of 96.3% for 12 combined ECG leads. The fact that different levels of accuracy are achieved for different ECG leads may suggest that the classifier takes into account something different from brain activity. An additional limitation of this approach lies in the analysis of ECG morphology, as it increases the likelihood of classifying patients based on the side effects and physiological impacts of administered psychiatric medications. Different classes of drugs prescribed for specific psychiatric disorders exert distinct effects on various aspects of ECG morphology [96]. This limitation is less pronounced when HRV or R-R intervals are analyzed, as this type of signal lacks information about electrical potentials or specific time-dependent events, although it is also susceptible to medication-induced changes [74]. Another issue comes from the fact that the ECG morphology of different leads depends on the shape of the thoracic chest [97]. In that context, schizophrenic patients are a distinct group when it comes to their metabolic functioning and, thus, the presented body type [98]. For these reasons, we consider classification based solely on ECG morphology to be inadequate for the psychiatric diagnosis. In addition, a complex 12-lead ECG signal requires more complex equipment than a single-lead Polar H10.

The above-mentioned example shows mainly that the acquisition of complex signals may not always be a desired solution to the given problem, particularly in a situation where we do not have direct access to the reasoning behind the given conclusion, as in the case of DL algorithms [84]. This could also be a problem for EEG whole brain resting state signals.

HRV and ERP address the described problem by isolating and extracting distinct functional activities from the base signal, reducing its complexity. Thus, these approaches may reduce the risk of inadequate data analysis and overfitting [92]. In the case of HRV, this extraction appears to be without disadvantages as the rest of the ECG signal outside of the R-R intervals is not an established significant information carrier of brain activity. This lack of disadvantages does not apply to ERP as it is extracted from an EEG data source that represents brain activity as a whole, resulting in a loss of potentially significant data.

Finally, when evaluating the EEG methodology in relation to ECG in psychiatry, it is crucial to note that both resting-state EEG and ERP reflect cortical activity [83], with the ERP focusing on smaller scale networks, such as the temporal–prefrontal network in the case of MMN [99]. In contrast, changes in HRV measured by the ECG represent the ANS activity regulated by deep brain structures [93, 100] and provide information on the affective states mediated by ANS activity [94]. Taking into account this information, ECG and EEG analysis do not overlap in their utility and could potentially be treated as complementary methods that provide different types of insight into brain activity.

### 4.6 Wearables in context of other disease monitoring approaches

In comparison to other diagnostic and treatment monitoring methods in healthcare, wearable devices tend to be the most cost-effective. This depends on the context of the disease we chose to monitor and the general availability of systemic healthcare [101]. In the case of ECG measurements with the use of Polar H10, at a price point lower than 100$, this tool offers a significantly more affordable option compared to other advanced diagnostic methods. In addition, it is less time-consuming and readily available with high reusability, without the need for highly specialized personnel for it to be operational. With the proper software, it is ready to use at home.

As wearables mostly rely on general physiological signals, they are not self-sufficient in their diagnostic and monitoring tasks, especially in psychiatric healthcare. They are a source of transdiagnostic information [102] and require contextual interpretation through software or a trained physician. But the same applies even to other more robust diagnostic methods, e.g. neuroimaging (MRI) [103].

A very important disease monitoring approach in medicine in general is laboratory blood testing, including genetic profiling. In the case of psychiatry, this method presents challenges in interpretation due to the absence of established diagnostic biomarkers and the high variability in genetically determined traits, complicating the integration of genetic tests into routine clinical practice [25].

This raises the question of the area of application of these methods. In contrast to the above-mentioned, wearable HRV monitoring is possibly much more publicly available. This allows it to be used outside of a clinical setting, for example, in rural areas and low-income countries, where the availability of professional healthcare is lacking as a first-line method of selective screening [104]. Another strategy would be wide use of the method for population screening for psychotic disorders that could enable a faster diagnosis of the disease in the prodromal stage, reducing the burden on systemic healthcare. As shown in [46], good quality monitoring can be achieved even in a non-standardized at-home setting, potentially broadening the application beyond healthcare facilities (e.g. for patients with limited mobility, living in a remote location, or under-home quarantine).

Another aspect is the monitoring of disease treatment and progression using telemedicine, as HRV is correlated with the severity of psychotic diseases [39]. This would enable the carrying out of treatment plans in more controlled ways as one of the main treatment problems is low compliance and the recurrence of symptoms due to the patient losing continuity of treatment and contact with professional healthcare [105]. It would also potentially allow for intervention before the aggravation of symptoms.

Such large-scale proceedings would not be possible using neuroimaging or EEG which are available primarily in specialized hospital settings, and not even laboratory testing.

### 4.7 Conclusions

In this study, we presented a classification approach, using short-duration R-R interval windows based on ECG signals, to differentiate between schizophrenia / bipolar disorder patients and healthy control individuals.

Our primary contribution lies in evaluating the accuracy of a classification-driven wearable-based diagnostic protocol. We examined the publicly available dataset containing samples from 60 persons. We implemented a state-of-the-art classification framework with a rigorous two-level cross-validation scheme for unbiased patient evaluation and automatic hyperparameter optimization. In addition, we compared several cutting-edge classification techniques, including deep learning approaches, to provide a comprehensive assessment.

Our method-independent approach, which yields an accuracy of 83% in 5-fold cross-validation and 80% in leave-one-out cross-validation, represents a significant step toward real-world clinical application. Our study indicates that wearable devices may be a cost-effective diagnostic tool, both for in- and outpatients. These promising results suggest that our framework could support future developments in diagnostic tools that integrate traditional symptom-based criteria with physiological markers, e.g. derived from ECG signals for more precise and reliable diagnoses. In addition, our experiments correspond to a real-world scenario, assuming almost an unsupervised environment.

Furthermore, biomarker-based diagnostics supported by machine learning offer a potential pathway to improve the predictive validity and early interventions for schizophrenia and bipolar disorder in their prodromal phase. Our study opens the door to self-monitoring of the patient, even when performing daily activities when the ECG is collected over a longer time period. In addition to early detection, this may provide the opportunity to monitor the development of the disease.

## Data Availability

Zenodo repository

https://zenodo.org/records/8171266

## Statements of ethical approval

All participants were informed and asked for written consent prior to admission to the experiment. The experimental procedure in the study received approval from the Bioethics Committee of the Medical University of Silesia, No.: BNW/NWN/0052/KB1/135/I/22/23.

## Data availability statement

All data used in experiments are available on Zenodo at link https://doi.org/10.5281/zenodo.8171266 while the source code for selected experiments is publicly available on a GitHub repository at https://github.com/iitis/Polar-HRV-classification.

